# Variation and Standardization in Prior Authorization Requirements

**DOI:** 10.1101/2025.02.07.25321895

**Authors:** Aya Zaari Jabri, Jacob Asher, Jacqueline Sandling, Kevin Schulman, David Scheinker

## Abstract

**Importance:** Prior authorization (PA) rules are neither regulated nor standardized.

**Objective:** To quantify the variation in PA rules of four US health insurers and examine the potential for standardization.

**Design:** Services and medications were identified using their Healthcare Common Procedure Coding System (HCPCS) codes. We manually examined PA rules to identify a simple set of categories applicable across insurers. We categorized HCPCS codes and their PA rules with large language models and verified the results manually. We identified the HCPCS codes for which all insurers require PA, use the same criteria to determine if PA is necessary, and have the same requirements to obtain PA.

**Setting:** Commercial insurance prior authorization rules from the provider manuals of Humana, United Healthcare, Anthem California, and Aetna.

**Participants:** In-network providers and internally managed contracts. Out-of-network providers and external PA management vendors were excluded.

**Results:** Across insurers, HCPCS codes were categorized as: medical and surgical; behavioral health; or special programs. To determine if PA is necessary, Anthem California provides a medical criteria document for each HCPCS code and the other insurers use 6 criteria. Anthem California has 18 requirements for prior authorization and the other insurers have 5 requirements. The number of HCPCS codes for which PA is necessary is 3,048 for Humana, 2,660 for United Healthcare, 2,188 for Anthem California, and 1,162 for Aetna. Of the 5,475 HCPCS codes for which PA was necessary with any insurer, 3% (157) were common to all insurers and 56.4% (3,088) were unique to one insurer. There were no HCPCS codes for which all insurers used the same criteria to determine if PA was necessary nor the same requirements to receive PA.

**Conclusions:** Across 4 US commercial insurers, there is significant variation in the HCPCS codes for which PA is necessary. A simple framework for categorizing HCPCS codes may facilitate the evaluation and standardization of PA rules.

**Key Points:** 

**Question:** Do prior authorization rules, purportedly derived from evidence-based coverage policies, differ across the commercial plans of four US insurers?

**Finding:** A simple framework was identified to reproduce the prior authorization rules of four commercial insurers. All four insurers required prior authorization for 3% (157 of 5,475) of HCPCS codes while for 56.4% (3,088 of 5,475) of HCPCS only one insurer required prior authorization.

**Meaning:** Prior authorization rules differed significantly across insurers. Using a common framework to define prior authorization rules may allow insurers to increase the concordance of evidence-based rules.

## Introduction

The United States health insurance market comprises 317,987 individual plans, approximately one plan for every 1,000 people in the country.^1^ The complexity associated with a lack of standardization in these contracts contributes at least $250 billion of the $400 billion in annual billing and administrative waste.^2,3^ Simplifying or standardizing this market could eliminate much of this waste, saving time for physicians, nurses, hospitals, and insurers.^1,4^ A comparative study of health systems in countries with hybrid public-private payer models suggests that this complexity is neither essential nor inevitable.^5^

Prior authorization (PA) is a process requiring physicians to obtain insurer approval for payment before providing care. It is intended to prevent low value care as part of the larger umbrella of utilization management. PA rules are neither standardized nor regulated and differ across insurers.^6^ Opportunities to improve PA exemplify the costs, care delays, and inefficiencies associated with administrative complexity. A care provider may have hundreds of PA requirements differing across hundreds of health plan contracts.^3^ Physicians surveyed by the American Medical Association report that PA is associated with delays in patient care, serious adverse events, and burnout.^7^ Patients report that PA causes delays in care, increased anxiety, and increased administrative burden.^8^ The American Medical Association, American Hospital Association, and America’s Health Insurance Plans support PA reform and the transition to electronic PA.^9–11^

While digital methods have been adopted for other financial transactions in healthcare, such as claim submissions and eligibility and benefits verification, prior authorization lags significantly behind. In 2022, 97% and 90% of medical plans reported using fully electronic claims submissions and eligibility and benefits verification, but only 28% reported using fully electronic PA.^12^ Adoption of electronic PA has been hindered by poor interoperability between health plans and providers, and the complexity that arises from the differences in PA requirements across plans. Although traditional Medicare does not require PA, relying primarily on retrospective denials, the rules for PA vary extensively across Medicare Advantage providers.^6^

The opportunity to improve healthcare contracts through standardization has been compared to successful similar efforts in other industries such as mortgages.^13^ Before 1971, states, trade associations, and banks had different mortgage laws, forms, and requirements. The complexity increased uncertainty, wait times, and costs for borrowers applying for mortgages. The complexity also hindered lenders seeking to syndicate mortgages on a secondary market.^13,14^ Fannie Mae and Freddie Mac, large government-sponsored buyers of mortgages, led the efforts to standardize mortgages and develop uniform mortgage instruments that significantly reduced variation, complexity, and costs of home mortgages. Uniform mortgages, processed primarily with algorithms instead of ad-hoc manual review, now account for over 90% of residential loans.^15^ Computable contracts, contracts designed for electronic interpretation and administration, may have the potential to provide similar benefits to health plan administration.^16^

In this study, we examine the potential for standardization by developing a simple framework applicable to the PA rules of four commercial US health insurerers and quantifying the variation in their PA rules.

## Methods

### Context

#### Provider manuals

Contracts between health plans and providers stipulate the terms and conditions of payment for services. Detailed instructions for specific transactions are typically included in publicly accessible provider manuals, which serve as appendices to health plan contracts. We downloaded the provider manuals for the commercial line of business from four US insurers: Humana, Aetna, United Healthcare, and Anthem (Details in Appendix 1).^17–22^ Anthem-Blue Cross, a division of Elevance, has separate provider manuals for each state. We selected the California manual since it is applicable to the largest patient population. To check for the sensitivity of the results to the inclusion of a California specific plan, we repeated the analysis with California omitted.

#### Prior Authorization

Prior authorization (PA) requires physicians to obtain insurer approval for payment before providing care, as a condition for reimbursement. PA requirements differ across insurers and across plans offered by each insurer. Depending on the structure of their insurance plans, each insurer’s provider manual contains numerous lists and criteria identifying when PA is necessary. Each PA list or criteria applies to some plans, locations, categories of services, or combinations thereof. We limited the scope of the work to the rules for in-network providers and internally managed contracts. We excluded rules for out-of-network providers, determining provider network status, and PA managed by external vendors.

#### Coding

Provider manuals identify services and medications with a variety of codes. The Current Procedural Terminology (CPT) is a coding system defined by the American Medical Association to describe medical, surgical, and diagnostic services provided by healthcare professionals. The Healthcare Common Procedure Coding System (HCPCS) is maintained by the Centers for Medicare and Medicaid Services and presents two levels of codes. The first level is equivalent to the CPT coding standards and the second level identifies services and medications that are not included in the first level. When HCPCS codes are not specific enough to identify an individual drug product in the PA process, the National Drug Code (NDC), a unique 11-digit number used and updated by the Food and Drug Administration, can be used to identify each medication in the US. Each insurer identifies services and medications with an HCPCS code (or equivalent CPT code) except United Healthcare, which uses brand and name for medications.

### Approach

#### Developing a standard framework

We manually examined the rules and logic of the provider manuals to identify simple categories of services across insurers. For each service and each provider manual, we identify the criteria to determine if PA is necessary (e.g., PA may be necessary depending on the age of the patient); when PA is necessary (e.g., PA is necessary for those under 18 years old); and the requirements to obtain PA (e.g., step therapy is required for the PA of a medication).

#### Extracting codes and PA rules into the framework

HCPCS codes were extracted from the insurer manuals and processed using large language models (LLMs). We prompted GPT, the LLM of OpenAI, to extract HCPCS codes from the unstructured text of each page and recorded those in a Python dictionary. Python is a popular programming language and a dictionary is a data structure well suited for examining and quantifying properties of the data stored. To ensure the accuracy of the extraction, we manually validated the output by comparing the number of HCPCS codes identified by the LLM to the number of services listed on the corresponding page of the provider manual. The details of the data structure, the underlying code, and the algorithms developed to examine the data are freely available upon reasonable request and in Appendix.

For each HCPCS code, we manually identified the criteria to determine if PA is necessary and the requirements to obtain PA (many HCPCS codes are listed with the same criteria to determine if PA is necessary and the requirements for PA).

#### Quantifying variation

Using the categories identified, we quantified similarities and differences in how services are categorized. We identified the HCPCS codes for which all insurers necessitate PA, use the same criteria to determine when PA is necessary, and have the same requirements for PA. Analogously, we identified the HCPCS codes for which exactly one, two, or three insurers: necessitate PA, use the same criteria to determine when PA is necessary, and have the same requirements for PA. We identified services and medications that are not explicitly identified by a HCPCS/CPT/NDC code or are not accompanied by unambiguous guidelines. We identified codes that appear more than once and with differing rules.

##### Sensitivity of the results to the inclusion of Anthem California

To test the sensitivity of the results to the inclusion of Anthem California, we repeated our analyses restricting to PA rules applicable to California. We reproduced the analyses identifying services for which PA is necessary across all insurers; the differences in criteria and requirements for PA; and the number of services for which PA is necessary by number of insurers.

## Results

### Framework for prior authorization rules

Across insurers, HCPCS codes were categorized as: medical and surgical; medication (restricted to medical benefit drugs); behavioral health; or special programs. Aetna’s provider manual used the category special programs for HCPCS codes with complex PA and, for other insurers, we used this category for services not identified by a HCPCS code and services for which the guidelines are not explicit and unambiguous.

Anthem California provides a medical document for each HCPCS code that includes when the code requires PA. For the other three insurers, the 6 criteria to determine if PA is necessary are: the state where care will be provided, the age of the patient, the diagnoses, additional services to be provided simultaneously, the site of service, and, for some prosthetics, orthotics, and durable medical equipment, whether the retail purchase or cumulative rental costs exceed a certain threshold (Details in Appendix 3). Anthem California has 18 categories of requirements for PA: photos, sleep study, X-rays and tracings, letters from mental health providers, visual fields, plastic surgery evaluation, psychiatric evaluation, medical clearance, history, physical and ultrasound evaluation, Haller Index +/-CT report, audiometry interpretation, nutritional consultation, BMI, documentation of participation in a weight management program, prosthetics evaluation, or MD prescription. The other three insurers have 5 categories of requirements for PA: site of service review, plan coverage review, dollar amount verification, working with a benefit management vendor or another process for a service category or provider network (we refer to this as Vendor or other process), and step therapy for medication (Details in Appendix 4).

The categorization of HCPCS codes, criteria to determine if PA is necessary, and the requirements for PA differ across insurers and for each insurer differ across categories. Aetna categorizes services and medications into medical and surgical; medications; behavioral health; and special programs; Humana does not use special programs category; United Healthcare includes behavioral health services in the medical and surgical category; and Anthem includes medications in the medical and surgical category and does not use special programs as a category (Figure 1).

**Figure 1:**
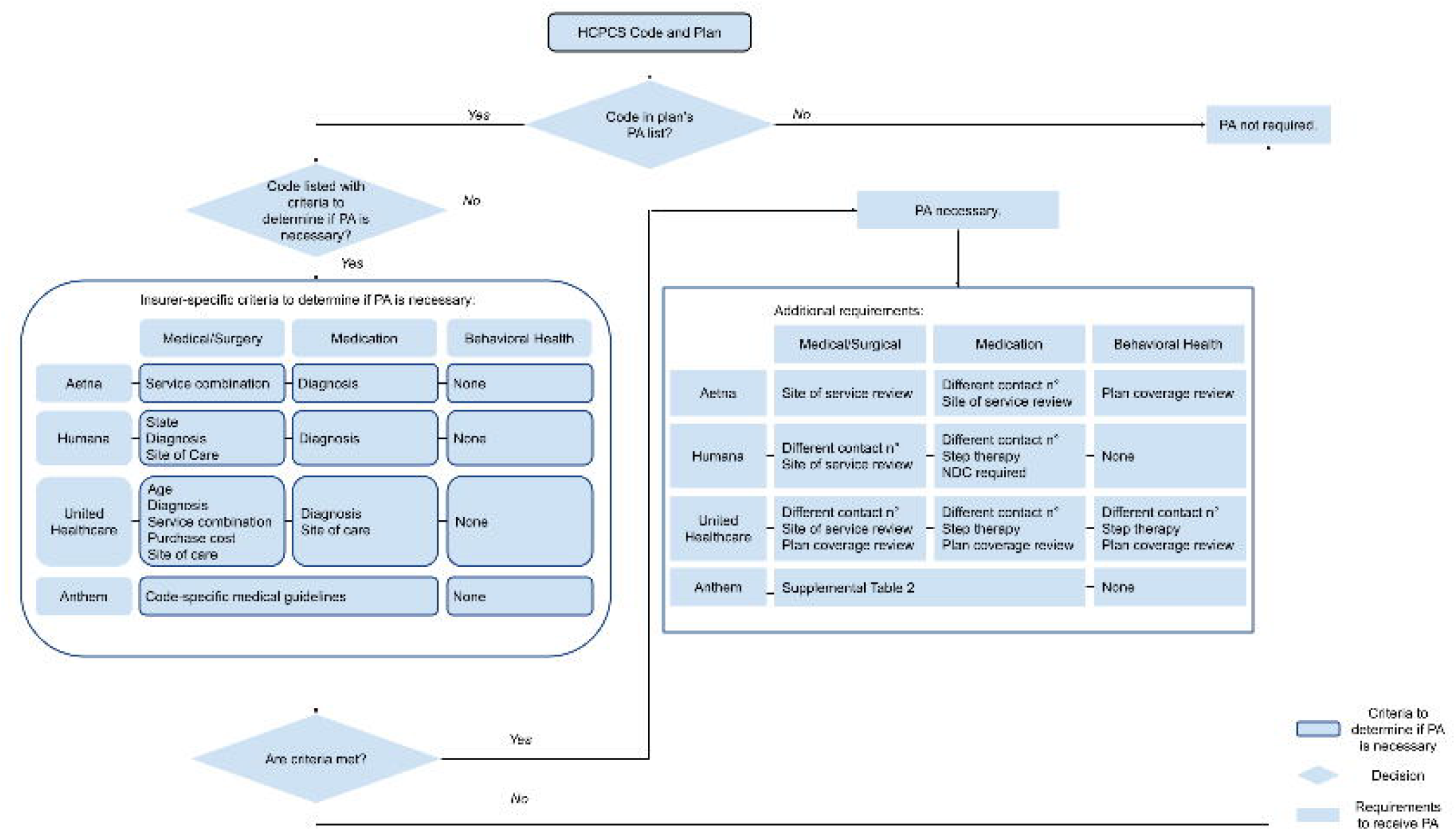

### Variation across insurers

#### Differences in the HCPCS codes for which PA is necessary

Prior authorization (PA) is necessary for 3,048 HCPCS codes for Humana, 2,660 for United Healthcare, 2,188 for Anthem California, and 1,162 for Aetna. Of the 5,475 HCPCS codes for which PA is necessary with any insurer, it is necessary for all insurers for 3% (157) of codes, necessary for three insurers for 16% (882) of codes, necessary for two insurers for 25% (1,348) of codes, and necessary for 1 insurer for 56.4% (3,088) of codes (Table 1). For each insurer, the Medical and Surgical category has the most HCPCS codes for which PA is necessary: 2,655 for Humana, 2,247 for United Healthcare, and 573 for Aetna. (Figure 2).

**Table 1:**
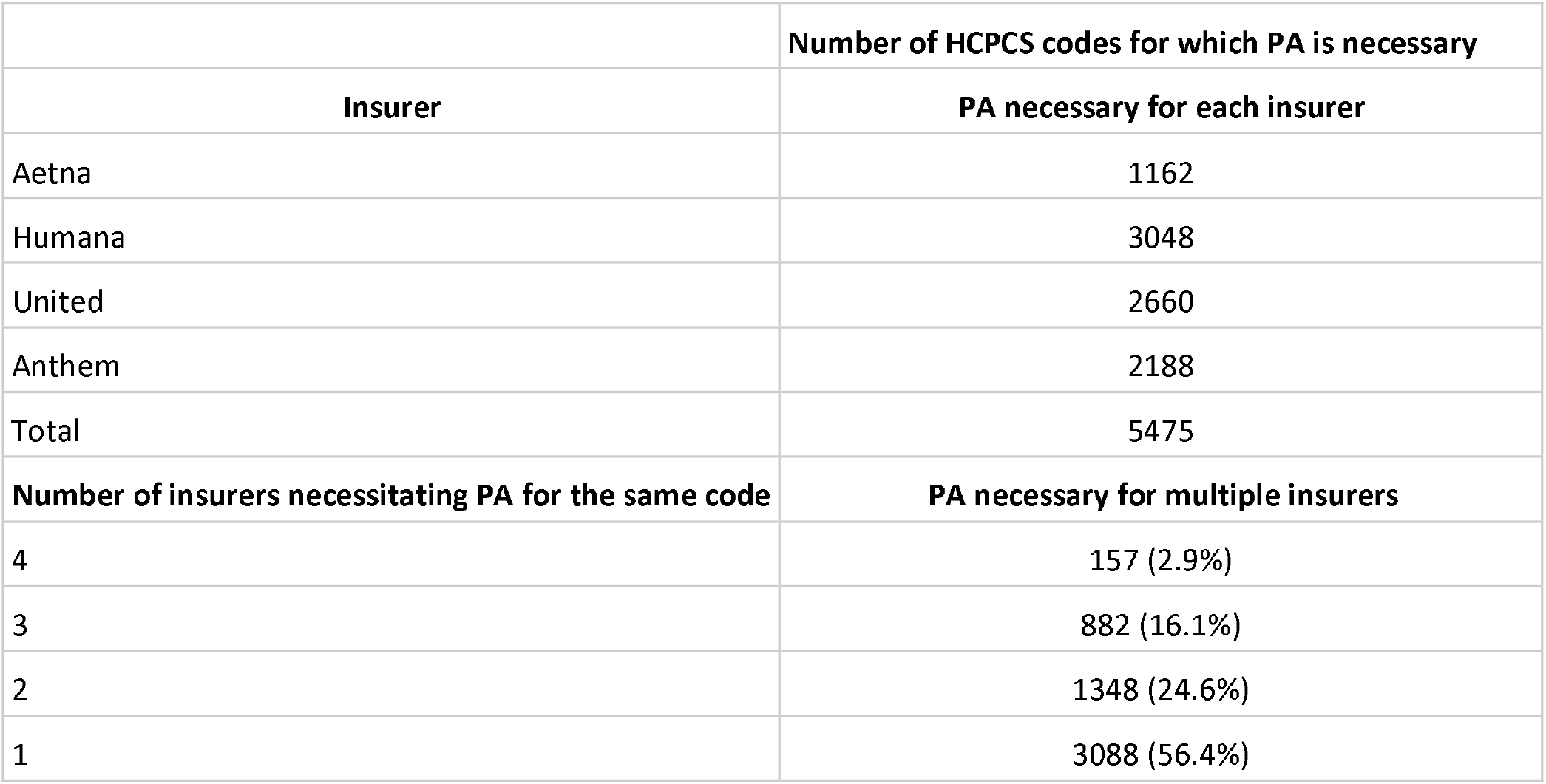
HCPCS codes for which PA is necessary for each insurer and for multiple insurers.

**Figure 2:**
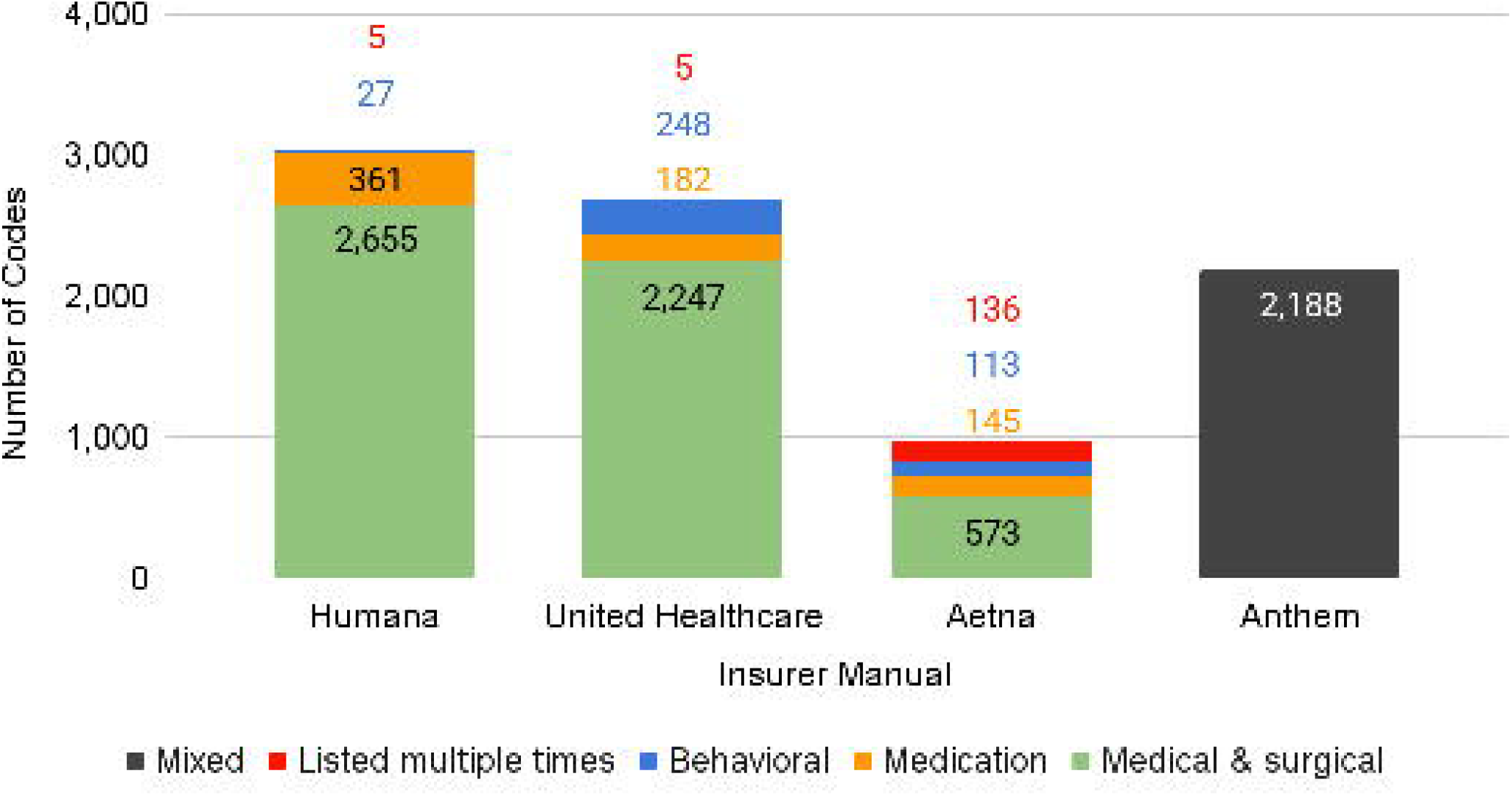

#### Variation in the classifications of HCPCS codes, criteria to determine if PA is necessary, and requirements for PA

There was significant variation across insurers in the frequency with which each criteria was used to determine if PA is necessary. The use of criteria to determine if PA is necessary varied by more than 10 fold across insurers: for medical and surgical services and medication, Aetna required at least one criteria to determine whether PA is necessary for 19 of 718 (2.6%) of codes; Humana, for 102 of 3026 (3.4%) of codes; UHC, for 1654 of 2407 (68.7%) of codes; and Anthem, for 217 of 2142 (10.1%) of codes (Table 2 and eTable 1). A site of service review is required for PA for 139 medical and surgical services and 91 medications by Aetna; 11 medical and surgical services by Humana; and 33 medical and surgical services by United Healthcare (Table 2 and eTable 1). HCPCS codes are categorized differently by insurers (eTable 2)

**Table 2:** Criteria to determine if prior authorization is necessary and requirements for prior authorization, for HCPCS codes in the categories Medical and Surgical and Medication

|  | Aetna |  | Humana |  | United Healthcare |  | Anthem |
| --- | --- | --- | --- | --- | --- | --- | --- |
|  | Medical and Surgical (n = 573) | Medication (n = 145) | Medical and Surgical (n = 2660) | Medication (n = 366) | Medical and Surgical (n = 2247) | Medication (n = 160) | Code-specific guidelines |
| **Criteria to determine if PA is required** |  |  |  |  |  |  |  |
| State | 0 | 0 | 4 | 0 | 858 | 0 | 0 |
| Age |  |  |  |  | 1 |  |  |
| Diagnosis | 0 | 15 | 87 | 2 | 287 | 9 | 0 |
| Service combination | 4 | 0 | 0 | 0 | 77 | 0 | 0 |
| Purchase cost |  |  |  |  | 243 |  |  |
| Site of care | 0 | 0 | 9 | 0 | 1039 | 9 | 0 |
| Code-specific medical guidelines | 0 | 0 | 0 | 0 | 0 | 0 | 217 |
| None | 569 | 130 | 2560 | 364 | 602 | 151 | 1925 |
| **Requirements for prior authorization** |  |  |  |  |  |  |  |
| Step therapy | 0 | 0 | 0 | 99 | 0 | 0 | 0 |
| NDC required | 0 | 0 | 0 | 58 | 0 | 0 | 0 |
| Site of service review | 139 | 91 | 11 | 0 | 33 | 0 | 0 |
| Plan coverage review | 0 | 0 | 0 | 0 | 22 | 150 | 0 |
| Vendor or other process* | 0 | 1 | 27 | 6 | 27 | 9 | 434 |
| All other requirements | 0 | 0 | 0 | 0 | 0 | 0 | eTable 1 |
| None | 434 | 54 | 2622 | 220 | 2165 | 1 | 1211 |

\*Prior authorization is managed by a benefit management vendor or there is another process for a service category or provider network

There were no HCPCS codes for which all insurers used the same criteria to determine if PA was necessary and 33 HCPCS codes for which two insurers had the same criteria to determine if PA was necessary. There were no HCPCS codes for which all insurers had the same requirements for PA, 4 HCPCS codes for which 3 insurers had the same requirements for PA, and 62 HCPCS codes for which two insurers had the same requirements for PA (Table 3).

**Table 3:** Number of insurers sharing the same criterion to determine if PA is necessary the same or requirements for PA for an HCPCS code

|  | Number of insurers sharing<br>the same criteria or requirement for<br>a given HCPCS code |  |  |  |  |
| --- | --- | --- | --- | --- | --- |
|  | 1 | 2 | 3 | 4 | Total HCPCS codes |
| **Criteria to determine if prior**<br>**authorization is necessary** |  |  |  |  |  |
| State | 862 | 0 | 0 | 0 | 862 (16%) |
| Age | 1 | 0 | 0 | 0 | 1 (0%) |
| Diagnosis | 358 | 21 | 0 | 0 | 379 (7%) |
| Service combination | 81 | 0 | 0 | 0 | 81 (1%) |
| Purchase cost | 243 | 0 | 0 | 0 | 243 (4%) |
| Site of care | 1108 | 12 | 0 | 0 | 1120 (20%) |
| **Requirement for prior**<br>**authorization** |  |  |  |  |  |
| Step therapy | 99 | 0 | 0 | 0 | 99 (2%) |
| NDC required | 58 | 0 | 0 | 0 | 58 (1%) |
| Site of service review | 436 | 4 | 0 | 0 | 440 (8%) |
| Plan coverage review | 169 | 0 | 0 | 0 | 169 (3%) |
| Vendor or other process* | 807 | 58 | 4 | 0 | 869 (16%) |

#### Sensitivity of the results to the inclusion of Anthem California

The results did not change significantly when the above analyses were restricted to PA rules pertaining to California (Appendix 6; eTables 3 – 6). The primary difference was in chiropractic therapy, where four services listed in the Humana manual required PA only in a subset of states (AZ, GA, IL, KY, OH, FL), but not in California.

## Discussion

We reproduced the prior authorization (PA) rules of the provider manuals or four commercial health plans, Humana, Aetna, United Healthcare, and Anthem California in a simple common framework with 4 categories of services, 6 types of criteria to determine if PA is necessary, and 23 types of requirements for PA. We identified the 5,489 HCPCS codes for which prior authorization is necessary: 3,048 for Humana, 2,660 for United Healthcare, 2,188 for Anthem, and 1,162 for Aetna. We quantified considerable variation across plans, including that all four insurers agreed that PA is necessary for only 4% of HCPCS codes. The variations identified suggest significant opportunities for standardization and the framework developed suggests a practical approach to standardizing.

Our findings of the variation across commercial plans in the services, for which PA is necessary and the requirements to receive PA, match the concerns of physicians about the PA process.^7,8^ There is broad consensus on the burdens and costs imposed by the PA process on patients, physicians, and insurers.^9–12,23^ There is far less consensus in the medical literature on how to simplify the process without creating unintended consequences such as increasing low value care or reducing insurer autonomy. The complexity of the PA process, and of health insurance contracts, is driven in part by regulatory differences across states highlighting the tension between national standardization and state autonomy. For example, Texas exempts physicians from an insurer’s PA requirements for a service if they receive approvals on at least 90% of their requests for that service during a six-month period.^24^ In contrast, Minnesota will soon prevent the use of prior authorization for the non-medication parts of cancer care and mental health care.^25^ A strength of the present work is that we explicitly capture differences across states, illustrating a method to reduce that source of complexity. The variation and inconsistencies in PA rules identified in this work offer a starting point for simplification and rationalization.

Our findings provide some data for why the PA process is so burdensome for physicians and patients. First, we found significant variability across insurers in services for which PA is necessary. This is consistent with the absence of rules requiring health plans to justify or provide data to support PA requirements. None of the logic in the provider manuals revealed why one plan determines that a given service requires PA while another does not. Second, we found variability within health plan provider manuals, such as which criteria are used to determine if PA is necessary and what the requirements are for PA. This is also consistent with the absence of any oversight or standards for health plan PA requirements. Our results support the proposition that standardized computable contracting frameworks have the potential to preserve the clinical rationale and content of the current system, while reducing the ambiguity and complexity required to administer health care payments. Subsequent research should use our framework to identify common services and medications for which PA requirements differ significantly across insurers and evaluate these differences against the available medical evidence.

As part of their charge to reduce administrative complexity and facilitate interoperability, a recently approved rule from Centers for Medicare & Medicaid Services (CMS) mandates payers to transition to electronic prior authorization by 2026. The rule includes standardized formats for computer-to-computer communication, such as Health Level 7 Fast Healthcare Interoperability Resources (FHiR) Prior Authorization application programming interface (API).^23^ However, the rule does not require insurers to standardize which services require PA nor the logic or documentation required for PA. Absent such standardization, the transition will digitize, rather than reduce the complexity and variability of the current process. Subsequent work should examine the specific mechanics of the most frequent reasons for denials, such as whether they are due to incomplete information in the original PA request or time pressures for payers to respond, and where standardization and algorithmic tools may be most helpful.

The United States Congress is considering bills to establish standardized PA requirements for Medicare Advantage plans.^26,27^ The framework used here would allow government agencies and health plans to analyze, justify, and reconcile the variation in and across providers. Such an approach could enable the creation of computable contracts to facilitate a standardized PA process. At a minimum, computable contracts would automatically identify duplications, ambiguities, and contradictions. Their ultimate promise is a uniform, evidenced-based foundation for electronic PA.

## Limitations

This work examined the PA requirements in the manuals of 4 providers of health insurance. The three insurers United Healthcare, Anthem, and Aetna are the three largest health insurers in the US commercial market. However, our analysis only examined the manual of Anthem California and not other states due to the structure of its manual. In addition, this work considers only the commercial line of business. Humana’s share of the US commercial market is less than 2%,^28^ and in 2023 Humana announced its intent to exit the Employer Group Commercial Medical Products business entirely.^29^ Although Humana’s manual for commercial plans will soon become defunct, its inclusion, as well as the inclusion of Anthem’s California manual, demonstrate that a simple framework reproduces the structures of plans of variable size and phases of the insurance plan product life cycle.

Our work focused on commercial plans, complementing research in the variation in PA rules in Medicare plans.^6^ We limited our focus to the rules for in-network providers and internally managed contracts, omitting other providers and provider network status and program structure (vendor-managed versus internally managed).

We relied on publicly available provider manuals and coverage policies from insurer websites, which were not linked to specific employer groups. These materials typically reflect default PA requirements applied to fully insured plans or serve as general guidance for providers. However, such plans operate under a distinct regulatory framework and may customize prior authorization requirements, though in practice these often align closely with the general guidance provided in standard manuals.

Effective operation of PA requires plans to maintain a significant managed care team including coverage policy committees and clinical reviewers that must withstand regulatory oversight and audits focused on process integrity. Internal health plan assessment of the business impact of PA must include tracking and trending denial overturns via appeals as well as the granular impact on actual claims paid out and, therefore, remains a complex, opaque accounting exercise that is reviewed annually. Balancing the frequent abrasiveness of the PA experience for both providers and patients with employer satisfaction and clinical cost savings remains a challenge.

We did not examine the clinical evidence for the PA requirements, a promising area for future work. Due to space constraints, the current work does not list the exact criteria to determine when PA is necessary nor the specific requirements for PA. For those interested in examining the results in more detail, for example, for which HCPCS codes Aetna does not require PA and other insurers do, an algorithm providing these for each HCPCS code is available upon reasonable request.”

The current analysis represents a single snapshot of the provider manuals, it is not dynamic, but PA requirements change over time. The present findings may be manually updated in response to published changes. Future work could integrate APIs to enable dynamic updates to provider rules. The resources required to develop our model were less than 0.000001% of the revenue of each of the insurers considered; the costs of expanding our approach to dynamically reproduce all provider manuals rules should not be prohibitive.

## Conclusion

The prior authorization rules of four commercial US insurance plans differed significantly but were compatible with a surprisingly simple framework of 6 criteria to determine if PA is necessary and 24 requirements to obtain PA. The significant variation identified in this work and the practical framework introduced may be a starting point for standardization efforts.

## Data Availability

All data and code produced in the present study are available upon reasonable request to the authors.

## Abbreviations

(PA): Prior Authorization
(CPT): Current Procedural Terminology
(HCPCS): Healthcare Common Procedure Coding System
(NDC): National Drug Code

## References

1. Tseng P, Kaplan RS, Richman BD, Shah MA, Schulman KA. Administrative Costs Associated With Physician Billing and Insurance-Related Activities at an Academic Health Care System. JAMA. 2018;319(7):691. doi:10.1001/jama.2017.19148

2. Sahni NR, Carrus B, Cutler DM. Administrative Simplification and the Potential for Saving a Quarter-Trillion Dollars in Health Care. JAMA. 2021;326(17):1677. doi:10.1001/jama.2021.17315

3. Shrank WH, Rogstad TL, Parekh N. Waste in the US Health Care System: Estimated Costs and Potential for Savings. JAMA. 2019;322(15):1501. doi:10.1001/jama.2019.13978

4. Scheinker D, Richman BD, Milstein A, Schulman KA. Reducing administrative costs in US health care: Assessing single payer and its alternatives. Health Serv Res. 2021;56(4):615–625. doi:10.1111/1475-6773.13649

5. Richman BD, Kaplan RS, Kohli J, et al. Billing And Insurance–Related Administrative Costs: A Cross-National Analysis: Study examines health care billing and insurance related administrative costs across several countries. Health Aff (Millwood). 2022;41(8):1098–1106. doi:10.1377/hlthaff.2022.00241

6. Gupta R, Fein J, Newhouse JP, Schwartz AL. Comparison of prior authorization across insurers: cross sectional evidence from Medicare Advantage. The BMJ. 2024;384:e077797. doi:10.1136/bmj-2023-077797

7. 2023 AMA Prior Authorization Physician Survey. American Medical Association; 2023. Accessed September 22, 2024. https://www.ama-assn.org/system/files/prior-authorization-survey.pdf

8. Chino F, Baez A, Elkins IB, Aviki EM, Ghazal LV, Thom B. The Patient Experience of Prior Authorization for Cancer Care. JAMA Netw Open. 2023;6(10):e2338182. doi:10.1001/jamanetworkopen.2023.38182

9. Reduce Practice Burdens with Electronic Prior Authorization. American Medical Association; 2017. Accessed September 22, 2024. https://www.ama-assn.org/sites/ama-assn.org/files/corp/media-browser/premium/arc/epa-promo-flyer.pdf

10. Prior Authorization. America’s Health Insurance Plans. 2024. Accessed September 22, 2024. https://www.ahip.org/issues/prior-authorization

11. Thompson A. AHA Urges CMS to Finalize the Improving Prior Authorization Processes Proposed Rule. Published online October 27, 2023. Accessed September 22, 2024. https://www.aha.org/lettercomment/2023-10-27-aha-urges-cms-finalize-improving-prior-authorization-processes-proposed-rule

12. 2023 CAQH Index Report. Council for Affordable Quality Healthcare, Inc; 2024. Accessed September 22, 2024. https://www.caqh.org/hubfs/43908627/drupal/2024-01/2023_CAQH_Index_Report.pdf

13. Zenios S, Favaro K. Precedent Thinking Method. Harv Bus Rev. Published online Forthcoming 2024.

14. Carrozzo P. Marketing The American Mortgage: The Emergency Home Finance Act Of 1970, Standardization And The Secondary Market Revolution. Real Prop Probate Trust J. 2005;39(4):765–805.

15. Randolph P. The Future of American Real Estate Law: Uniform Foreclosure Laws and Uniform Land Security Interest Act. Nova Law Rev. 1996;20(3):1113.

16. Istvan, Brooke, Nielsen Jr, Perry, Eluhu, Megan, et al. Applying Precedents Thinking to the Intractable Problem of Transaction Costs in Healthcare. Health Manag Policy Innov. 2024;9(3).

17. Participating provider behavioral health precertification list for Aetna. Published online August 1, 2024. Accessed April 4, 2025. https://www.aetna.com/content/dam/aetna/pdfs/aetnacom/healthcare-professionals/documents-forms/bh_precert_list.pdf

18. Participating provider precertification list for Aetna. Published online November 1, 2024. Accessed April 4, 2025. https://www.aetna.com/content/dam/aetna/pdfs/aetnacom/healthcare-professionals/2024_Precert_List.pdf

19. Humana Commercial Medication Preauthorization List. Published online January 8, 2025. Accessed April 4, 2025. https://docushare-web.apps.external.pioneer.humana.com/Marketing/docushare-app?file=5395286

20. Humana Commercial Preauthorization and Notification List. Published online June 11, 2024. Accessed April 4, 2025. https://docushare-web.apps.external.pioneer.humana.com/Marketing/docushare-app?file=5361564

21. Prior Authorization Requirements for UnitedHealthcare. Published online May 1, 2024. Accessed April 4, 2025. https://www.uhcprovider.com/content/dam/provider/docs/public/prior-auth/pa-requirements/commercial/UHC-Commercial-Advance-Notification-PA-Requirements-5-1-2024.pdf

22. Anthem Local PPO Precertification/Prior Authorization List. Published online November 22, 2024. Accessed April 4, 2025. https://www.anthem.com/docs/public/inline/CA_PPO_PA_List.pdf

23. Centers for Medicare & Medicaid Services. Medicare and Medicaid Programs; Patient Protection and Affordable Care Act; Advancing Interoperability and Improving Prior Authorization Processes for Medicare Advantage Organizations, Medicaid Managed Care Plans, State Medicaid Agencies, Children’s Health Insurance Program (CHIP) Agencies and CHIP Managed Care Entities, Issuers of Qualified Health Plans on the Federally-Facilitated Exchanges, Merit-Based Incentive Payment System (MIPS) Eligible Clinicians, and Eligible Hospitals and Critical Access Hospitals in the Medicare Promoting Interoperability Program. Fed Regist. 2024;89(42):8762–8763.

24. TMA Seeks Prior Authorization Reform This Legislative Session. Accessed April 4, 2025. https://www.texmed.org/TexasMedicineDetail.aspx?Pageid=46106&id=65351

25. 10 states have tackled prior authorization so far in 2024. American Medical Association. August 19, 2024. Accessed April 4, 2025. https://www.ama-assn.org/practice-management/prior-authorization/10-states-have-tackled-prior-authorization-so-far-2024

26. Rep. Kelly, Mike [R-PA-16]. Improving Seniors’ Timely Access to Care Act of 2024.; 2024. Accessed January 11, 2025. https://www.congress.gov/bill/118th-congress/house-bill/8702/text

27. Sen. Whitehouse S [D R. Text -S.5612 - 118th Congress (2023-2024): Prior Authorization Relief Act. December 19, 2024. Accessed April 4, 2025. https://www.congress.gov/bill/118th-congress/senate-bill/5612/text

28. AMA identifies market leaders in health insurance. American Medical Association. December 12, 2023. Accessed April 4, 2025. https://www.ama-assn.org/press-center/press-releases/ama-identifies-market-leaders-health-insurance

29. Humana to Exit Employer Group Commercial Medical Products Business. Accessed April 4, 2025. https://press.humana.com/news/news-details/2023/Humana-to-Exit-Employer-Group-Commercial-Medical-Products-Business/default.aspx

