## Supplemental Materials for "Variation and Standardization in Prior Authorization Requirements"

Insights from a Framework for US Commercial Insurers

Scheinker

**Contents:**

**Appendix 1 - Data sources**

**Appendix 2 - Algorithm and database development**

**Appendix 3 - Criteria to determine if prior authorization is necessary**

**Appendix 4 - Requirements to obtain prior authorization**

eTable 1: Anthem California requirements for PA

**Appendix 5 - Categories of HCPCS codes**

eTable 2: Variation in HCPCS code categories within and across insurers

**Appendix 6 - Sensitivity of the results to the inclusion of Anthem California**

eTable 3: HCPCS codes for which PA is necessary for each insurer and for multiple insurers, restricting to PA rules for California.

eTable 4: Criteria to determine if prior authorization is necessary and requirements for prior authorization, for HCPCS codes in the categories Medical and Surgical and Medication, restricting to PA rules for California

eTable 5: *Number of insurers sharing the same criterion to determine if PA is necessary for an HCPCS code*

eTable 6: *Number of insurers sharing the same requirements for PA for an HCPCS code*

**Appendix 1 - Data sources**

We used the prior authorization lists provided in the provider’s manuals of the 4 insurance companies. The lists correspond to the 2024 PA policy and are publicly available through the following links:

- Aetna:

The study scope includes the following plans: Aetna commercial plans, Banner Aetna plans, Sutter health Aetna plans, Texas Health Aetna plans and excludes students plans and Allina plans from the special programs count of services.

- Medical/Surgery, Medication and special programs: <https://www.aetna.com/content/dam/aetna/pdfs/aetnacom/healthcare-professionals/2024_Precert_List.pdf>
- Behavioral health: <https://www.aetna.com/content/dam/aetna/pdfs/aetnacom/healthcare-professionals/documents-forms/bh_precert_list.pdf>
- Humana:
  - Medical/Surgery and behavioral health: <https://docushare-web.apps.external.pioneer.humana.com/Marketing/docushare-app?file=5361564>
  - Medication: <https://docushare-web.apps.external.pioneer.humana.com/Marketing/docushare-app?file=5395286>
- United Healthcare:
  - Medical/Surgery, Medication and behavioral health: <https://www.uhcprovider.com/content/dam/provider/docs/public/prior-auth/pa-requirements/commercial/UHC-Commercial-Advance-Notification-PA-Requirements-5-1-2024.pdf>
- Anthem:

We include all the services that are reviewed by Anthem and reviewed by Anthem or Carelon.

<https://www.anthem.com/docs/public/inline/CA_PPO_PA_List.pdf>

**Appendix 2 – Algorithm and database development**

The first provider’s manual that we analyzed is Aetna’s manual. Four categories of services are explicitly mentioned and were therefore utilized for other manuals: Medical/Surgery, Medication, Behavioral health, and Special programs. United healthcare provides a PA list combining all categories and a second list providing additional medications. The second list does not use HCPCS codes, thus a lookup table was designed to capture the additional drugs.

The database is a set of python dictionaries (key-value) built as follows: the keys of the dictionary are HCPCS codes. For each HCPCS code, several subdictionaries correspond to the plan and category (e.g. Humana medication), constituting the key of the subdictionary. The value of each subdictionary is a list of elements defining the inputs and outputs. For instance, the first element of the list corresponds to the state of service. If, for a specific plan, the service only requires prior authorization in California, the first element of the list is ‘CA’. The algorithm is then able to process the requests for all services across all plans and service categories in a single and standardized approach.

The interactive algorithm was developed by defining a comprehensive list of the information that the insurer requires for prior authorization and splitting it into input and output elements. Inputs are necessary specifications to determine if a service requires a prior authorization. Outputs are provided to the user specifying additional requirements to receive prior authorization.

**Appendix 3 – Criteria to determine if prior authorization is necessary**

**Anthem California:**

**Medical guidelines**

Insurers utilize established medical guidelines and criteria as benchmarks to assess the medical necessity of a requested service. Those guidelines are designed and reviewed by insurers and provided as referring documents. Anthem Californian provides, for each service mentioned in the provider list, a medical guidelines reference document that indicates the conditions under which a prior authorization may be required.

**Humana, United Healthcare, and Aetna:**

**State where care is provided**

The geographic location in which medical care is administered is a factor in the approval process as state-specific regulations and policies can influence coverage determinations. For instance, Humana requires prior authorization for chiropractic therapy only in Arizona, Georgia, Illinois, Kentucky, Ohio and South Florida.

**Age of the patient**

Patient age is a criterion in determining eligibility for certain medical treatments or services. For example, United Healthcare requires prior authorization for patient aged 18 or older for under percutaneous transcatheter closure procedures.

**Diagnosis**

The medical diagnosis plays a role in evaluating the appropriateness of a requested treatment or service as it ensures that the intervention corresponds to the documented medical necessity and greed upon by the insurer. For instance, Aetna requires prior authorization for Bortezomib only if multiple myeloma is diagnosed.

**Additional services to be provided simultaneously**

Concurrent services that are scheduled to accompany the primary treatment can be reviewed to ensure clinical compatibility and care planning. For instance, Aetna requires prior authorization for generators, neurostimulators (implantable), non-rechargeable C1767 when it is used with a list of services among which permanent percutaneous epidural implantation of the neurostimulator electrode array.

**Site of service**

The location where the service is delivered, such as a hospital, outpatient facility, or home setting, is another determinant in the approval process. This factor ensures that the site is both clinically appropriate and cost-effective. For example, Humana requires prior authorization for epidural injections only if provided in an outpatient setting.

**Retail purchase cost or cumulative rental**

For prosthetics, orthotics, and durable medical equipment, the cost—whether as a single purchase or cumulative rental—may be evaluated to determine if it exceeds a predetermined threshold. This assessment ensures that the request aligns with cost-effectiveness and necessity considerations. For example, United Healthcare requires prior authorization only for DME codes listed with a retail purchase or cumulative rental cost of more than $1,000.

**Appendix 4 - Requirements to obtain prior authorization**

**eTable 1: Anthem California requirements for PA**

| **Requirements to obtain PA** | **Number of codes** |
| --- | --- |
| Plastics evaluation | 111 |
| X-rays and tracings | 47 |
| Sleep study | 25 |
| Psych Eval | 23 |
| Visual fields | 7 |
| Mental health letter | 111 |
| Photos | 97 |
| Dollar amount verification | 222 |
| Medical clearance | 23 |
| Prosthetics evaluation | 222 |
| Nutritional consultation | 23 |
| History and physical and ultrasound evaluation | 5 |
| 2 mental health letters | 63 |
| Physician prescription | 222 |
| Haller Index +/- CT report | 3 |
| Evidence of 6-month weight management program | 23 |
| Body mass index | 23 |
| Audiometry interpretation | 3 |

**Humana, United Healthcare, and Aetna:**

**Change in the contact numbers to submit the request**

For certain services and medications, the approval process may require a the use of different contact numbers for submission. This happens, for example, when the code is managed by a benefit management company or when different processes apply to specific service categories or provider networks.

**Step therapy**

A step therapy can also be required as part of the precertification process, inviting the patients to try alternative treatments before more expensive or risky ones. For instance, Humana requires a step therapy through a company preferred drug for some drugs such as the immune globulin.

**National Drug Code request**

Concerning the medication category, the insurance company can request the submission of the National Drug Code (NDC) instead of the HCPCS code as part of the precertification process for a higher level of accuracy. For example, Humana’s list mentions that “All shared HCPCS codes and not otherwise classified codes require a corresponding National Drug Code to be billed on all claims”.

**Site of service review**

The insurance company can also require a site of service review as this can impact the patient experience, accessibility and cost of care of the healthcare service. For instance, in Aetna’s preauthorization list, cochlear devices are subject to the medical necessity review of the site of service for commercial members.

**Plan coverage review**

A plan coverage review may be required as part of the prior authorization process to ensure that the specific healthcare service is covered under the patient’s plan and determines the level of coverage. For instance, United Healthcare mentions that a plan coverage review is necessary for the bariatric surgery services as they are not covered by some benefit plans.

**Appendix 5 - Categories of HCPCS codes**

Of the 113 services categorized by Aetna as behavioral, 10 are categorized as medical and surgical by Humana, 49 by United Healthcare, and 73 by Anthem.

**eTable 2: Variation in HCPCS code categories within and across insurers**

|  |  | Aetna | | | | Humana | | | United Healthcare | | | Anthem | |
| --- | --- | --- | --- | --- | --- | --- | --- | --- | --- | --- | --- | --- | --- |
|  |  | Medication | Medical/  Surgical | Behavioral | Special programs | Medication | Medical/  Surgical | Behavioral | Medical/  Surgical | Medication | Special programs | Medical/  Surgical/  Medication | Behavioral |
| Aetna | Medication | 145 |  |  |  | 124 |  |  |  | 95 |  | 3 |  |
|  | Medical/  Surgical |  | 573 | 90 | 46 |  | 399 |  | 370 |  |  | 297 |  |
|  | Behavioral |  |  | 113 |  |  | 10 | 13 | 49 |  |  | 73 | 10 |
|  | Special programs |  |  |  | 467 | 4 | 377 |  | 142 |  | 197 | 77 |  |
| Humana | Medication |  |  |  |  | 366 | 5 |  | 10 | 143 |  | 10 |  |
|  | Medical  /Surgical |  |  |  |  |  | 2660 |  | 899 |  | 181 | 1056 |  |
|  | Behavioral |  |  |  |  |  |  | 27 |  |  |  |  | 14 |
| United Healthcare | Medical/ Surgical |  |  |  |  |  |  |  | 2247 | 3 |  | 588 |  |
|  | Medication |  |  |  |  |  |  |  |  | 160 |  | 8 |  |
|  | Special Programs |  |  |  |  |  |  |  |  |  | 256 | 17 |  |
| Anthem | Medical/  Surgical/  Medication |  |  |  |  |  |  |  |  |  |  | 2142 |  |
|  | Behavioral |  |  |  |  |  |  |  |  |  |  |  | 46 |

**Appendix 6 - Sensitivity of the results to the inclusion of Anthem California**

eTable 3: HCPCS codes for which PA is necessary for each insurer and for multiple insurers, restricting to PA rules for California.

|  | **Number of HCPCS codes for which PA is necessary** |
| --- | --- |
| **Insurer** | **PA necessary for each insurer** |
| Aetna | 1162 |
| Humana | 3044 |
| United | 2660 |
| Anthem | 2188 |
| Total | **5471** |
| **Number of insurers necessitating PA for the same code** | **PA necessary for multiple insurers** |
| 4 | 157 (2.9%) |
| 3 | 882 (16.1%) |
| 2 | 1348 (24.6%) |
| 1 | 3084 (56.4%) |

eTable 4: Criteria to determine if prior authorization is necessary and requirements for prior authorization, for HCPCS codes in the categories Medical and Surgical and Medication, restricting to PA rules for California

|  | Aetna | | Humana | | United Healthcare | | Anthem |
| --- | --- | --- | --- | --- | --- | --- | --- |
|  | Med/surg | Medication | Med/surg | Medication | Med/surg | Medication | Med/surg & medication |
| Total | 573 | 145 | 2656 | 366 | 2247 | 160 | 2142 |
| Criteria to determine whether PA is required |  |  |  |  |  |  |  |
| State |  |  |  |  | 858 |  |  |
| Age |  |  |  |  | 1 |  |  |
| Diagnosis |  | 15 | 87 | 2 | 287 | 9 |  |
| Service combination | 4 |  |  |  | 77 |  |  |
| Purchase cost |  |  |  |  | 243 |  |  |
| Site of care |  |  | 9 |  | 1039 | 9 |  |
| Code-specific medical guidelines |  |  |  |  |  |  | 217 |
| None | 569 | 130 | 2560 | 364 | 602 | 151 | 1925 |
| Requirements to obtain prior authorization |  |  |  |  |  |  |  |
| Different contact n° |  | 1 | 27 | 6 | 27 | 9 | 434 |
| Step therapy |  |  |  | 99 |  |  |  |
| NDC required |  |  |  | 58 |  |  |  |
| Site of service review | 139 | 91 | 11 |  | 33 |  |  |
| Plan coverage review |  |  |  |  | 22 | 150 |  |
| All other requirements |  |  |  |  |  |  | eTable 2 |
| None | 434 | 54 | 2618 | 220 | 2165 | 1 | 1211 |

*Prior authorization is managed by a benefit management vendor or there is another process for a service category or provider network

eTable 5: *Number of insurers sharing the same criterion to determine if PA is necessary for an HCPCS code*

|  | Number of insurers sharing  the same criteria for a given code | | | | |  |  |  |  |  |
| --- | --- | --- | --- | --- | --- | --- | --- | --- | --- | --- |
|  | 0 | 1 | 2 | 3 | 4 | total services  requesting the criteria | | | total services | share of services requesting the criteria |
| State | 4613 | 858 | 0 | 0 | 0 | 858 | | | **5471** | 16% |
| Age | 5470 | 1 | 0 | 0 | 0 | 1 | | | **5471** | 0% |
| Diagnosis | 5092 | 358 | 21 | 0 | 0 | 379 | | | **5471** | 7% |
| Service combination | 5390 | 81 | 0 | 0 | 0 | 81 | | | **5471** | 1% |
| Purchase cost | 5228 | 243 | 0 | 0 | 0 | 243 | | | **5471** | 4% |
| Site of care | 4351 | 1108 | 12 | 0 | 0 | 1120 | | | **5471** | 20% |

eTable 6: *Number of insurers sharing the same requirements for PA for an HCPCS code*

|  | Number of insurers sharing  the same criteria for a given code | | | | |  |  |  |  |  |
| --- | --- | --- | --- | --- | --- | --- | --- | --- | --- | --- |
|  | 0 | 1 | 2 | 3 | 4 | total services  requesting the criteria | | | total services | share of services requesting the requirement |
| different contact n° | 4602 | 807 | 58 | 4 | 0 | 869 | | | **5471** | 16% |
| Step therapy | 5372 | 99 | 0 | 0 | 0 | 99 | | | **5471** | 2% |
| NDC required | 5413 | 58 | 0 | 0 | 0 | 58 | | | **5471** | 1% |
| Site of service review | 5031 | 436 | 4 | 0 | 0 | 440 | | | **5471** | 8% |
| Plan coverage review | 5302 | 169 | 0 | 0 | 0 | 169 | | | **5471** | 3% |
